# ARTIFICIAL INTELLIGENCE IN PERCUTANEOUS CORONARY INTERVENTION: IMPROVED PREDICTION OF PCI-RELATED COMPLICATIONS USING AN ARTIFICIAL NEURAL NETWORK

**DOI:** 10.1101/2020.08.17.20177055

**Authors:** Hemant Kulkarni, Amit P. Amin

## Abstract

**Importance:** Complications after percutaneous coronary intervention (PCI) are common and costly. Risk models for predicting the likelihood of acute kidney injury (AKI), bleeding, stroke and death are limited by accuracy and inability to use non-linear relationships among predictors. Additionally, if non-linear relationships among predictors can be leveraged, then the prediction of any adverse event (i.e. “the patient who will not do well with PCI”) is perhaps of greater interest to clinicians than prediction of adverse events in isolation.

**Objective:** To develop and validate a set of artificial neural networks (ANN) models to predict five adverse outcomes after PCI – AKI, bleeding, stroke, death and one or more of these four (‘any adverse outcome’).

**Design:** Cross-sectional study, using institutional NCDR CathPCI data.

**Setting and participants:** 28,005 patients undergoing PCI at five hospitals in the Barnes-Jewish Hospital system.

**Main Outcome(s):** AKI, bleeding, stroke, death, and one or more of these four (‘any adverse outcome’). We divided 28,005 PCI patients into a training cohort of 21,004 (75%) and a test cohort of 7,001 (25%). We used an artificial neural network (ANN) multilayer perceptron (MLP) model to predict each outcome based on a set of 278 encoded and preprocessed variables. Model accuracy was tested using area under the receiver-operating-characteristic curve (AUC). Performance and validation of the MLP model was compared with existing regression models using integrated discrimination improvement (IDI) and continuous net reclassification index (NRI).

**Results:** The prevalence of AKI, bleeding, stroke and death in the study cohort was 4.6%, 3.6%, 0.3% and 1.1%, respectively. The fully trained MLP model achieved convergence quickly (< 10 epochs) and could predict accurately predict AKI (77.9%), bleeding (86.5%), death (90.3%) and any adverse outcome (80.6%) in the independent test set. However, prediction of stroke was not satisfactory (69.9%). Compared to the currently used models for AKI, bleeding and death prediction, our models showed a significantly higher AUC (range 1.6% – 5.6%), IDI (range 4.9% –7.2%) and NRI (range 0.07 – 0.61).

**Conclusions and Relevance:** By using neural network-based models, we accurately predict major adverse events after PCI. Larger studies for replicability and longitudinal studies for evidence of impact are needed to establish these artificial intelligence methods in current PCI practice.

## INTRODUCTION

Percutaneous coronary intervention (PCI) is common in the United States, performed in ∼600,000 PCI procedures annually.(1) With an increasing pressure on hospitals to improve the quality and value of their services, reducing the costs of elective PCI, is an important opportunity to explore. In fact, alternative payment models such as the Centers for Medicare and Medicaid Services (CMS) episode payment models (EPMs), commonly known as “bundled payments”, are accelerating hospitals to prepare for the shift in reimbursement from ‘payment for volume’ to ‘payment for value’.(2)

In this context, risk prediction models for adverse outcomes in patients undergoing PCI have an important role to play in the practice of ‘value-driven PCI’. Complications after PCI such as bleeding, or acute kidney injury (AKI) or stroke or death, after PCI are not considered reliably predictable. Although validated risk-prediction models based on regression methods, can quantify a patient’s risk of these adverse events,(3–6) they have not been considered highly accurate. Furthermore, adverse events rarely occur in isolation. The link between bleeding and AKI after PCI is well known.(7) Similarly, the association between bleeding and mortality, and AKI and mortality are also well established.(8,9) Ultimately, as clinicians taking care of patients, the goal of prediction models is not to identify events in isolation, but to accurately predict the entire spectrum of adverse events that are of importance to patients and providers alike.

Conceptually, another challenge that underlies these predictions is the existence of nonlinear relationships among traditional risk factors and outcomes, and the impact of these nonlinear associations on mortality, the ultimate adverse event.(10) Traditional regression modelling is unable to handle these complex relationships. Artificial intelligence (Al) methods, such as artificial neural networks (ANN), are particularly suited to tackle the challenges of scalability and high dimensionality of data with complex relationships among predictors and show promise in the field of PCI outcomes predictions. Our main motivation was to develop an ANN-based system of models that can simultaneously predict the risk of major complications in PCI patients using the same set of pre-procedural and procedural characteristics as predictors. Using a large dataset of PCI patients enrolled at a multi-hospital system, we developed and validated ANN-based risk-stratification system to be used as an aid to decision making during PCI.

## METHODS

### Study Population

This study used NCDR CathPCI Registry data (1) for the Barnes-Jewish Corporation (BJC) Healthcare hospitals spanning a period from 1 July 2009 to 30 April 2018. The BJC Healthcare hospitals include the following seven hospitals – Alton Memorial Hospital, Alton, IL; Barnes Jewish Hospital, St Louis, MO; Barnes Jewish St. Peters Hospital, St. Peters, MO; Boone Hospital Center, Columbia, MO; Christian Hospital, St Louis, MO; Missouri Baptist Medical Center, St. Louis, MO; and Progress West Healthcare, O’Fallon, MO. During this period a total of 30,520 PCIs conducted at these hospitals from which we included 28,005 (91.8%) in this study. This study was approved by Washington University’s institutional review board, and no informed consent was required.

### Predictors and Outcomes

This study included five outcomes and 100 predictors. The predictors represented following categories of information: baseline clinical variables (n = 31), admission related variables (n = 9), stress studies (n = 4), cardiac catheterization findings and imaging studies (n = 16), medications (n = 6), laboratory investigations (n = 5), PCI-characteristics at the start of PCI (n = 10) and PCI procedure-related variables (n = 19). The NCDR Cath registry evaluates characteristics, treatments, and outcomes of patients undergoing PCI and/or diagnostic catheterization.(1) Full definitions for all predefined variables are available at the American College of Cardiology’s NCDR website (http://cvquality.acc.org/en/NCDR-Home/Registries/Hospital-Registries.aspx). Details of the variables included in the study are also provided in Table 1. Our machine-learning algorithm attempted to predict from these predictors one of the following five outcomes during index hospitalizations – AKI, bleeding, stroke, death and a composite outcome that represented the occurrence of any one of these four outcomes. Our aim was to simultaneously predict a spectrum of four most common and severe PCI-related adverse outcomes using a multiclass prediction. However, since some of the combinations of these four outcomes were very rare (Supplementary Table 1), we did not have adequate representation of such classes. Alternatively, we proceeded with development of different models (using the same set of inputs) to predict each of the four outcomes separately and to predict a fifth composite outcome that represented a combination of the four outcomes (hereinafter referred to as “any adverse outcome”).

Definitions of these outcomes were the same as that used in the NCDR CathPCI registry as follows: AKI: Acute Kidney Injury Network (AKIN) stage 1 or greater or a new requirement for dialysis following PCI (11); Bleeding: any ONE of the following: 1. Bleeding event w/in 72 hours; OR 2. Hemorrhagic stroke; OR 3. Tamponade; OR 4. Post-PCI transfusion for patients with a pre-procedure hemoglobin > 8 g/dL and pre-procedure hemoglobin not missing; OR 5. Absolute hemoglobin decrease from pre-PCI to post-PCI of ≥ 3 g/dl AND pre-procedure hemoglobin ≤ 16 g/dL AND pre-procedure hemoglobin not missing (12); Stroke: transient ischemic attack, hemorrhagic stroke OR ischemic stroke; Death: death during index hospitalization.

### Machine-learning modeling approach

We used a feed-forward, artificial neural network framework designed as a multi-layer perceptron (MLP) for all predictive modeling in this study. The detailed approach to data preparation, preprocessing, model specification, training and validation are depicted in Figure 1. The first step in data preparation included the splitting of a randomly shuffled dataset into a derivation set (n = 21,004 – 75% of the entire dataset) and a validation dataset (n = 7,001 – 25% of the entire dataset). This split was conducted only once and all training for machine-learning algorithms used data from the derivation set while all models were validated on data from the validation set.

**Figure 1.**
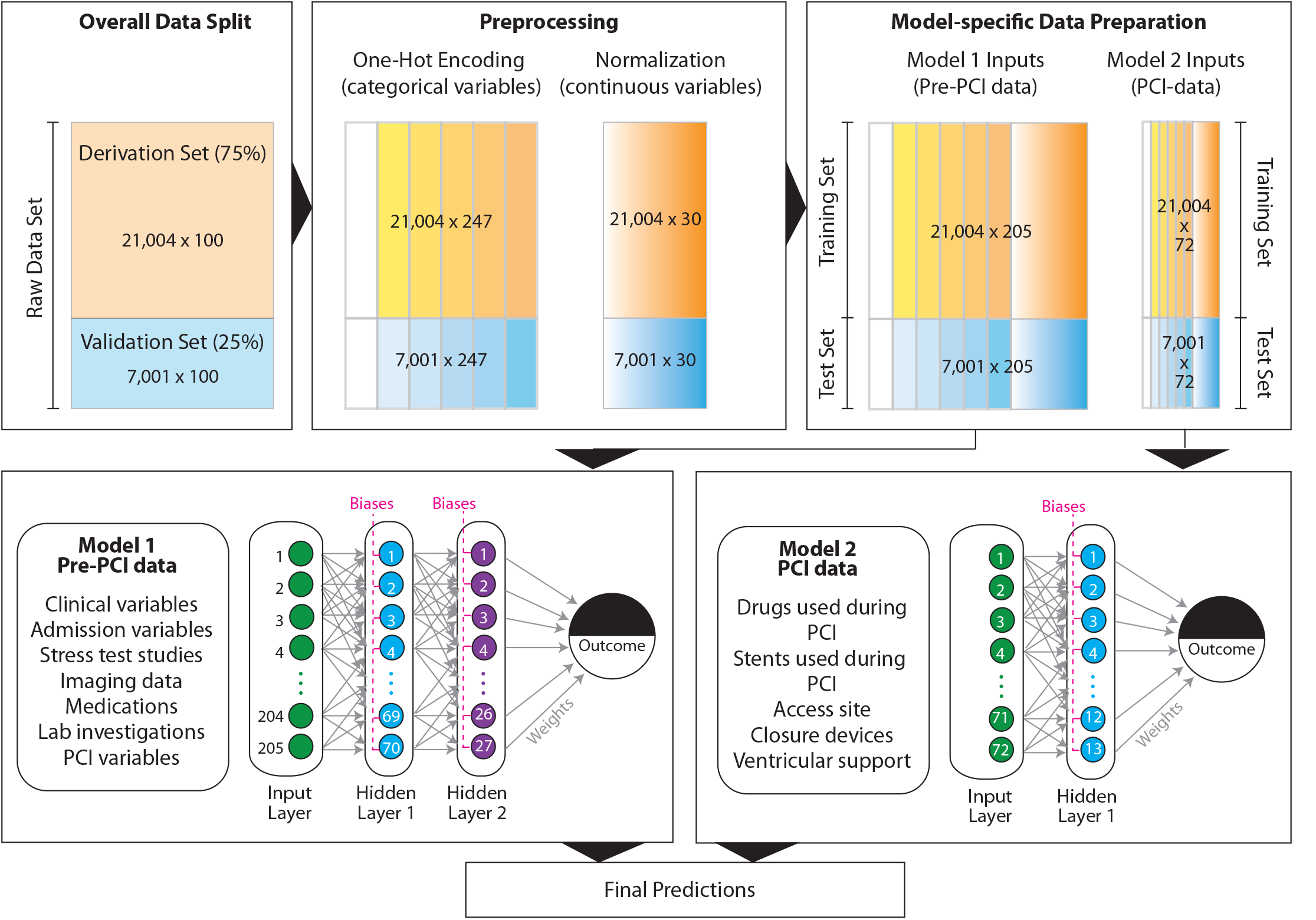
Analysis pipeline for MLP-based predictive model. For details, please see text. The derivation and training sets are color coded as orange while the validation and test sets are color coded blue. Categorical variables are indicated by a stepwise gradient and continuous variables are indicated by a continuous gradient.

The second step included data preprocessing using variable encoding. We aimed to maximize the information contained within a variable and therefore did not discard any records with missing values. Rather, we coded all missing values for all variables as –1 to include missing information as a separate category. Categorical variables (whether nominal or ordinal) were coded using a one-hot (13) encoding approach such that a vector (instead of a single scalar value) was used to denote the categorical value for each patient. The one-hot vector was encoded as 1 (for the category observed) or 0 (otherwise).(13) Continuous variables were standard-normalized as 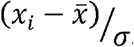, where Xi is the i^th^ observation, 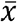 is the mean and σ is the standard-deviation.

Our third step in the analytical pipeline included a generation of two datasets that were used as inputs to two different learning models for each outcome. From a clinical standpoint, the estimation of pre-PCI probability of an outcome is important and valuable for the PCI operator to make decisions about the PCI procedure. Therefore, we constructed Model 1 that incorporated all the baseline and pre-PCI variables (205 encoded variables) while Model 2 was constructed to include variables directly related to the PCI procedure (73 encoded variables). Predictions from these two models were then finally combined into a single prediction using logistic regression.

### Model architecture, specification and tuning

All the models contained an input layer (the encoded dataset, hereafter referred to as the training set), a variable number of hidden layers and an output layer (represented by the dichotomous outcome in question). As shown in Figure 1, the MLP model included estimations of the weights (indicated by gray arrows) and biases (indicated by magenta dashed line that is common to all neurons within a single hidden layer) using the standard back-propagation gradient estimation algorithm. All neurons in the hidden function used the sigmoid activation function. Final output of the MLP was the predicted probability for the adverse outcome in each patient. The number of hidden layers, the number of neurons within each hidden layer, the learning rate and the number of training epochs required for convergence were treated as hyperparameters and tuned using a grid search that provided optimum model performance.

The cost function used to optimize the model was cross-categorical entropy. Avoidance of overfitting was ensured using the following approach. For each training epoch, the estimated best fitting model was independently applied to the test set (the encoded dataset obtained from the validation set) to trace the classification accuracy. Model training continued as long as there was improvement in the classification accuracy for both the training and the independently assessed test set. If the model only showed accuracy improvement in the training set but showed a decreased accuracy for the test set, then a potential overfitting was interpreted, and model training was stopped.

### Statistical analysis

Descriptive statistics included mean and standard deviation for continuous variables and percentages for categorical variables. Significant difference in the distribution of the study variables across the Derivation and Validation sets was tested using Student’s t test (for continuous variables) or chi-square test (for categorical variables). Correction for multiple testing was done using the Bonferroni method. Importance of each variable used in the final MLP model was estimated using Olden’s algorithm.(14) Calibration plots were used to examine the overall predictive performance of the MLP by plotting the observed and expected risk deciles and fitting a lowess smoother to the data. Accuracy of classification was assessed by plotting a receiver-operating characteristic (ROC) and estimating the area under the ROC curve (AUC). Improvement in discrimination and reclassification was assessed using the Integrated Discrimination Improvement (IDI) and the net reclassification index (NRI). Statistical significance was tested at a type I error rate of 0.05.

All analyses were carried out using R (CRAN) or Stata (Stata Corp, College Station, Texas). Following programs were used for specific aspects of analyses: one-hot encoding (R package onehot (13)), normalization of continuous variables and MLP implementation (R package RSNNS (15)), variable importance estimation (R package NeuralNetTools (16)), calibration plots (pmcalplot (17) package in Stata), AUC comparison (roccomp command in Stata that uses the DeLong test to compare AUCs (18)), and IDI and NRI estimation (idi and nri packages, Michael Lunt, University of Manchester, Manchester, United Kingdom).

## RESULTS

### Study populationn

This study was based on 28,005 PCIs on 26,784 patients, most of whom (n = 25,589, 95.5%) underwent one PCI, 1,169 (4.4%) had two PCIs and 26 (0.1%) had three PCIs during the study period. The average age of the patients was 65.6 years. Obesity (body mass index > 30 kg/m^2^, 46.6%), diabetes mellitus (40.4%), dyslipidemia (84.5%), and hypertension (83.8%) chronic renal (26.4%) and chronic lung (17.6%) disease were the commonest comorbidities. The average pre-PCI left ventricular ejection fraction was 52.1%, and a large proportion (varying between 21%-47%) patients had a significant history of heart failure, coronary artery disease, myocardial infarction, peripheral artery disease, or previous cardiac interventions (PCI and/or coronary artery bypass graft(CABG)). This full dataset was randomly split into a derivation set (n = 21,004) and a validation set (n = 7,001) which were statistically comparable with respect to the 100 predictor variables included in the study (Table 1).

**Table 1.** Baseline characteristics, PCI-related data and observed outcomes in the study population

| Characteristic | Categories | Code | Training Set |  | Test Set |  | p |
| --- | --- | --- | --- | --- | --- | --- | --- |
|  |  |  | Mean / N | SD / % | Mean / N | SD / % |  |
| **Clinical characteristics** |  |  |  |  |  |  |  |
| Age (y) | --- | None | 65.6 | 12.05 | 65.48 | 12.28 | 0.463 |
| Female gender | Yes | 1 | 7041 | 33.52 | 2249 | 32.12 | 0.0314 |
| White Race | Yes | 1 | 18414 | 87.67 | 6140 | 87.7 | 0.9425 |
| Body Mass Index (Kg/m<sup>2</sup>) |  | None | 30.47 | 6.69 | 30.49 | 6.63 | 0.8049 |
| Current smoker |  | 1 | 5407 | 25.74 | 1818 | 25.97 | 0.7094 |
| Diabetes |  | 1 | 8510 | 40.52 | 2802 | 40.02 | 0.4664 |
| Hypertension |  | 1 | 17619 | 83.88 | 5853 | 83.6 | 0.5795 |
| Dyslipidemia |  | 1 | 17762 | 84.57 | 5896 | 84.22 | 0.4808 |
| Diabetes therapy |  | 1 | 307 | 1.46 | 105 | 1.5 | 0.349 |
|  | Diet | 2 | 562 | 2.68 | 211 | 3.01 |  |
|  | Oral | 3 | 3884 | 18.49 | 1305 | 18.64 |  |
|  | Insulin | 4 | 3731 | 17.76 | 1173 | 16.75 |  |
|  | Other | 5 | 22 | 0.1 | 8 | 0.11 |  |
| Chronic lung disease |  | 1 | 3702 | 17.63 | 1240 | 17.71 | 0.8693 |
| Chronic kidney disease |  | 1 | 5365 | 26.61 | 1742 | 25.88 | 0.2392 |
| Current dialysis |  | 1 | 643 | 3.06 | 220 | 3.14 | 0.7353 |
| Anemia |  | 1 | 1389 | 6.61 | 469 | 6.7 | 0.8022 |
| Family history of CAD |  | 1 | 5508 | 26.23 | 1825 | 26.07 | 0.7941 |
| Past history of myocardial infarction |  | 1 | 7004 | 33.35 | 2327 | 33.24 | 0.8682 |
| Past history of heart failure |  | 1 | 4434 | 21.11 | 1523 | 21.75 | 0.2543 |
| Past history of peripheral arterial disease |  | 1 | 3235 | 15.4 | 1021 | 14.58 | 0.0986 |
| Past history of valve surgery |  | 1 | 487 | 2.32 | 164 | 2.34 | 0.9076 |
| Past history of PCI |  | 1 | 10004 | 47.63 | 3289 | 46.98 | 0.3456 |
| Past history of CABG |  | 1 | 4511 | 21.48 | 1508 | 21.54 | 0.9116 |
| Past history of cerebrovascular disease |  | 1 | 3365 | 16.02 | 1102 | 15.74 | 0.5793 |
| Past history of heart failure within 2 weeks |  | 1 | 4724 | 22.49 | 1550 | 22.14 | 0.5415 |
| Cardiogenic shock within past 24 hours |  | 1 | 402 | 1.91 | 139 | 1.99 | 0.7066 |
| Cardiac arrest within past 24 hours |  | 1 | 304 | 1.45 | 104 | 1.49 | 0.8175 |
| NYHA class within past 2 weeks | I | 1 | 326 | 6.91 | 105 | 6.77 | 0.8959 |
|  | II | 2 | 1034 | 21.9 | 342 | 22.06 | 0.8959 |
|  | III | 3 | 1823 | 38.61 | 584 | 37.68 | 0.8959 |
|  | IV | 4 | 1538 | 32.58 | 519 | 33.48 | 0.8959 |
| Past history of other major surgery |  | 1 | 332 | 1.58 | 104 | 1.49 | 0.5767 |
| Time elapsed since last CABG (days) |  | None | 4227.66 | 2689.13 | 4217.65 | 2747 | 0.901 |
| Time elapsed since last PCI (days) |  | None | 1598.18 | 1811.28 | 1605.91 | 1830.02 | 0.8323 |
| Time since onset of symptoms (days) |  | 0 | 18992 | 90.08 | 6297 | 89.94 | 0.8813 |
|  |  | 1 | 1356 | 21.79 | 456 | 21.88 |  |
|  |  | 2 | 328 | 5.27 | 117 | 5.61 |  |
|  |  | 3 | 223 | 3.58 | 65 | 3.12 |  |
|  |  | 4 | 91 | 1.46 | 35 | 1.68 |  |
|  |  | 5 | 38 | 0.61 | 16 | 0.77 |  |
|  |  | 6 | 38 | 0.61 | 11 | 0.53 |  |
|  |  | 7 | 8 | 0.13 | 4 | 0.19 |  |
| Anginal classification within 2 weeks | None | 1 | 1254 | 5.97 | 403 | 5.76 | 0.3681 |
|  | CCS I | 2 | 759 | 3.61 | 287 | 4.1 |  |
|  | CCS II | 3 | 1859 | 8.85 | 599 | 8.56 |  |
|  | CCS III | 4 | 8130 | 38.71 | 2720 | 38.85 |  |
|  | CCS IV | 5 | 9002 | 42.86 | 2992 | 42.74 |  |
| Cardiomyopathy or LV dysfunction |  | 1 | 2859 | 13.61 | 1004 | 14.34 | 0.1262 |
| **Admission characteristics** |  |  |  |  |  |  |  |
| Insurance | Medicare/Medicaid only | 1 | 6982 | 33.24 | 2243 | 32.04 | 0.1064 |
|  | Multiple | 2 | 12730 | 60.61 | 4296 | 61.36 | 0.1064 |
| CAD Presentation | No symptom, no angina | 1 | 1004 | 4.78 | 324 | 4.63 | 0.666 |
|  | Symptom unlikely to be ischemia | 2 | 436 | 2.08 | 163 | 2.33 |  |
|  | Stable angina | 3 | 2553 | 12.15 | 853 | 12.18 |  |
|  | Unstable angina | 4 | 10515 | 50.06 | 3481 | 49.72 |  |
|  | NSTEMI | 5 | 4087 | 19.46 | 1400 | 20 |  |
|  | STEMI or equivalent | 6 | 2409 | 11.47 | 780 | 11.14 |  |
| Hospital status | Outpatient | 1 | 6082 | 28.96 | 2028 | 28.97 | 0.6108 |
|  | Outpatient converted to inpatient | 2 | 3952 | 18.82 | 1353 | 19.33 |  |
|  | Inpatient | 3 | 10968 | 52.22 | 3620 | 51.71 |  |
| Admit source | Emergency department | 1 | 5792 | 27.58 | 1965 | 28.07 | 0.7317 |
|  | Transfer from another acute care facility | 2 | 4418 | 21.04 | 1464 | 20.91 |  |
|  | Other | 3 | 10788 | 51.38 | 3571 | 51.01 |  |
| PCI status | Elective | 1 | 8025 | 38.21 | 2716 | 38.81 | 0.8336 |
|  | Urgent | 2 | 10084 | 48.01 | 3333 | 47.63 |  |
|  | Emergency | 3 | 2860 | 13.62 | 937 | 13.39 |  |
|  | Salvage | 4 | 35 | 0.17 | 12 | 0.17 |  |
| Inpatient | Inpatient for this episode | 1 | 14920 | 71.03 | 4973 | 71.03 | 0.9982 |
| Door to balloon time (min) |  | None | 77.45 | 336.78 | 65.81 | 82.56 | 0.3654 |
| Symptom action time (min) |  | None | 386.66 | 684.12 | 422.6 | 889.4 | 0.6099 |
| **Stress studies** |  |  |  |  |  |  |  |
| Stress echocardiogram | Low risk | 1 | 166 | 14.12 | 57 | 14.92 | 0.6096 |
|  | Intermediate risk | 2 | 941 | 80.02 | 298 | 78.01 |  |
|  | High risk | 3 | 67 | 5.7 | 27 | 7.07 |  |
|  | Unavailable | 4 | 2 | 0.17 | 0 | 0 |  |
| SPECT stress test | Low risk | 1 | 425 | 8.56 | 128 | 7.78 | 0.099 |
|  | Intermediate risk | 2 | 4357 | 87.74 | 1462 | 88.82 |  |
|  | High risk | 3 | 156 | 3.14 | 40 | 2.43 |  |
|  | Unavailable | 4 | 28 | 0.56 | 16 | 0.97 |  |
| Exercise stress test | Low risk | 1 | 43 | 12.36 | 11 | 10.28 | 0.4438 |
|  | Intermediate risk | 2 | 274 | 78.74 | 91 | 85.05 |  |
|  | High risk | 3 | 27 | 7.76 | 4 | 3.74 |  |
|  | Unavailable | 4 | 4 | 1.15 | 1 | 0.93 |  |
| Stress test with CMR | Low risk | 1 | 2 | 7.14 | 1 | 14.29 | 0.779 |
|  | Intermediate risk | 2 | 23 | 82.14 | 6 | 85.71 |  |
|  | High risk | 3 | 2 | 7.14 | 0 | 0 |  |
|  | Unavailable | 4 | 1 | 3.57 | 0 | 0 |  |
| **Imaging studies** |  |  |  |  |  |  |  |
| Coronary calcium score | Yes | 1 | 51 | 0.78 | 21 | 0.98 | 0.3871 |
| Calcium score |  | None | 856.61 | 961.73 | 609 | 841.03 | 0.3074 |
| Cardiac CTA | No disease | 1 | 18 | 28.13 | 11 | 42.31 | 0.1651 |
|  | 1 vessel disease | 2 | 17 | 26.56 | 3 | 11.54 |  |
|  | 2 vessel disease | 3 | 21 | 32.81 | 9 | 34.62 |  |
|  | 3 vessel disease | 4 | 2 | 3.13 | 3 | 11.54 |  |
|  | Indeterminate | 5 | 5 | 7.81 | 0 | 0 |  |
| Left main stenosis (%) |  | None | 12.3 | 23.99 | 12.38 | 24.18 | 0.8251 |
| Proximal LAD stenosis (%) |  | None | 40.41 | 39.57 | 40.11 | 39.74 | 0.615 |
| Mid/Distal LAD stenosis (%) |  | None | 58.45 | 37.79 | 58.72 | 37.61 | 0.6218 |
| Circumflex artery stenosis (%) |  | None | 54.09 | 39.51 | 53.54 | 39.48 | 0.3404 |
| Ramus stenosis (%) |  | None | 16.9 | 32.24 | 17.46 | 32.48 | 0.4474 |
| RCA stenosis (%) |  | None | 64.44 | 37.32 | 64.33 | 37.41 | 0.827 |
| Proximal LAD graft stenosis (%) |  | None | 14.33 | 32.74 | 13.75 | 32.06 | 0.727 |
| Mid/Distal LAD graft stenosis (%) |  | None | 25.3 | 40.37 | 25.89 | 40.68 | 0.6671 |
| Circumflex artery graft stenosis (%) |  | None | 48.06 | 46.09 | 50.72 | 46.44 | 0.1149 |
| RCA graft stenosis (%) |  | None | 51.26 | 45.86 | 52.78 | 45.93 | 0.3667 |
| Ramus graft stenosis (%) |  | None | 10.53 | 28.18 | 11.87 | 30.51 | 0.412 |
| Dominance | Left | 1 | 1702 | 8.11 | 550 | 7.86 | 0.5951 |
|  | Right | 2 | 17668 | 84.23 | 5927 | 84.74 |  |
|  | Co-dominant | 3 | 1605 | 7.65 | 517 | 7.39 |  |
| LV ejection fraction |  | None | 52.2 | 13.48 | 51.83 | 13.49 | 0.0867 |
| **Medications** |  |  |  |  |  |  |  |
| Antianginal - beta-blockers |  | 1 | 13541 | 87.32 | 4460 | 87.02 | 0.5799 |
| Antianginal - Ca-channel blockers |  | 1 | 4388 | 28.3 | 1437 | 28.04 | 0.7222 |
| Antianginal - Long acting nitrates |  | 1 | 4616 | 29.77 | 1540 | 30.05 | 0.7025 |
| Antianginal - ranolazine |  | 1 | 631 | 4.07 | 198 | 3.86 | 0.5156 |
| Antianginal - other |  | 1 | 740 | 4.77 | 254 | 4.96 | 0.5937 |
| Thrombolytics |  | 1 | 82 | 3.42 | 25 | 3.21 | 0.7774 |
| **Laboratory investigations** |  |  |  |  |  |  |  |
| Pre-PCI CKMB |  | None | 21.67 | 56.96 | 23.32 | 60.64 | 0.443 |
| Pre-PCI Tnl |  | None | 5.07 | 25.23 | 5.01 | 18.02 | 0.9118 |
| Pre-PCI TnT |  | None | 0.45 | 2.18 | 0.44 | 1.39 | 0.8698 |
| Pre-PCI Serum Creatinine |  | None | 1.22 | 1.15 | 1.22 | 1.18 | 0.9977 |
| Pre-PCI Hemoglobin |  | None | 13.19 | 2 | 13.21 | 2.03 | 0.6364 |
| **PCI variables before start of PCI** |  |  |  |  |  |  |  |
| Time of PCI start | 00:00 - 05:59 AM | 0 | 463 | 2.2 | 168 | 2.4 | 0.1027 |
|  | 06:00 AM - 11:59 AM | 1 | 10205 | 48.59 | 3385 | 48.35 |  |
|  | 12:00 Noon - 5:59 PM | 2 | 9102 | 43.33 | 3086 | 44.08 |  |
|  | 6:00 PM - 11:59 PM | 3 | 1234 | 5.88 | 362 | 5.17 |  |
| Day of PCI | Sunday | 0 | 600 | 2.86 | 204 | 2.91 | 0.5308 |
|  | Monday | 1 | 4164 | 19.82 | 1428 | 20.4 |  |
|  | Tuesday | 2 | 4117 | 19.6 | 1321 | 18.87 |  |
|  | Wednesday | 3 | 3830 | 18.23 | 1239 | 17.7 |  |
|  | Thursday | 4 | 3854 | 18.35 | 1316 | 18.8 |  |
|  | Friday | 5 | 3753 | 17.87 | 1280 | 18.28 |  |
|  | Saturday | 6 | 686 | 3.27 | 213 | 3.04 |  |
| Diagnostic catheterization done |  | 1 | 13971 | 66.52 | 4605 | 65.78 | 0.2568 |
| Other procedure with diagnostic catheterization |  | 1 | 2911 | 13.86 | 963 | 13.76 | 0.8271 |
| Fluoroscopy time (min) |  | None | 17.26 | 13.41 | 17.49 | 13.52 | 0.2189 |
| Fluoroscopy dose (mGy) |  | None | 2156.1 | 1494.94 | 2220.65 | 1447.97 | 0.4495 |
| Contrast volume (ml) |  | None | 186.67 | 84.11 | 187.17 | 83.4 | 0.665 |
| Number of diseased vessels |  | 0 | 265 | 1.26 | 73 | 1.04 | 0.2583 |
|  |  | 1 | 10097 | 48.07 | 3411 | 48.72 |  |
|  |  | 2 | 6290 | 29.95 | 2117 | 30.24 |  |
|  |  | 3 | 4352 | 20.72 | 1400 | 20 |  |
| Estimated glomerular filtration rate |  | None | 69.94 | 20.1 | 70.4 | 20.11 | 0.1016 |
| Cardiogenic shock at the start of PCI |  | 1 | 450 | 2.14 | 133 | 1.9 | 0.2176 |
| **PCI procedure related characteristics** |  |  |  |  |  |  |  |
| Glycoprotein IIb/IIIa inhibitors | Given | 1 | 2883 | 13.73 | 1015 | 14.5 | 0.1067 |
| Fondaparinux |  | 1 | 11 | 0.05 | 6 | 0.09 | 0.4522 |
|  |  | 2 | 2 | 0.01 | 2 | 0.03 |  |
|  |  | 3 | 1 | 0 | 0 | 0 |  |
| Low molecular weight heparin |  | 1 | 1789 | 8.52 | 574 | 8.2 | 0.5252 |
|  |  | 2 | 3 | 0.01 | 2 | 0.03 |  |
| Unfractionated heparin | Given | 1 | 10054 | 47.87 | 3352 | 47.88 | 0.9916 |
| Aspirin |  | 1 | 19935 | 94.92 | 6621 | 94.6 | 0.5628 |
|  |  | 2 | 117 | 0.56 | 43 | 0.61 | 0.5628 |
| Bivalirudin |  | 1 | 13532 | 64.43 | 4525 | 64.63 | 0.753 |
| Clopidogrel |  | 1 | 15517 | 73.88 | 5238 | 74.82 | 0.2914 |
|  |  | 2 | 70 | 0.33 | 21 | 0.3 |  |
| Ticlopidine |  | 1 | 20 | 0.1 | 8 | 0.11 | 0.8961 |
|  |  | 2 | 35 | 0.17 | 11 | 0.16 |  |
| Prasugrel |  | 1 | 2102 | 10.22 | 663 | 9.7 | 0.4622 |
|  |  | 2 | 40 | 0.19 | 13 | 0.19 |  |
| Ticagrelor |  | 1 | 2937 | 18.19 | 960 | 17.85 | 0.8284 |
|  |  | 2 | 42 | 0.26 | 12 | 0.22 |  |
|  |  | 3 | 1 | 0.01 | 0 | 0 |  |
| Number of drug eluting stents |  | None | 1.45 | 1.03 | 1.47 | 1.04 | 0.2011 |
| Number of bare metal stents |  | None | 0.18 | 0.56 | 0.18 | 0.57 | 0.4689 |
| Minimum stent diameter (mm) |  | None | 2.87 | 0.51 | 2.87 | 0.51 | 0.2165 |
| Total stent length (mm) |  | None | 31.48 | 21.93 | 31.98 | 22.23 | 0.1129 |
| Number of lesions |  | None | 0.13 | 0.6 | 0.13 | 0.61 | 0.6337 |
| Transradial access |  | 1 | 2460 | 11.71 | 799 | 11.41 | 0.4987 |
| Vascular closure device |  | 1 | 11595 | 55.2 | 3890 | 55.56 | 0.6001 |
| Intra-aortic balloon pump | Present at the start of PCI | 1 | 69 | 27.82 | 30 | 36.59 | 0.3239 |
|  | Inserted during PCI | 2 | 59 | 23.79 | 17 | 20.73 |  |
|  | Inserted after PCI | 3 | 120 | 48.39 | 35 | 42.68 |  |
| Other mechanical ventricular support | Present at the start of PCI | 1 | 50 | 26.74 | 14 | 25 | 0.4269 |
|  | Inserted during PCI | 2 | 116 | 62.03 | 32 | 57.14 |  |
|  | Inserted after PCI | 3 | 21 | 11.23 | 10 | 17.86 |  |
| **Outcomes** |  |  |  |  |  |  |  |
| Acute kidney injury | Observed | 1 | 955 | 4.55 | 328 | 4.69 | 0.6317 |
| Bleeding | Observed | 1 | 744 | 3.54 | 272 | 3.89 | 0.1838 |
| Stroke | Observed | 1 | 56 | 0.27 | 19 | 0.27 | 0.9466 |
| Death | Observed | 1 | 232 | 1.1 | 67 | 0.96 | 0.2982 |
| At least one adverse outcome | Observed | 1 | 1630 | 7.76 | 552 | 7.88 | 0.7371 |

### Classification using multi-layer perceptron models

Using hyperparameter grid search, we found that an MLP with two hidden layers provided optimum classification performance for Model 1 with respect to all the outcomes. The optimum number of neurons in the first and second hidden layers was 70 and 27, respectively. The results of model performance are described in Table 2, Figure 1 and Supplementary Figure 1.

**Table 2.** Predictive accuracy of the MLP classifier

| **Model** | **Outcome** | **Epochs** | **Accuracy in training set**<br>**AUC (95% CI)** | **Accuracy in test set**<br>**AUC (95% CI)** |
| --- | --- | --- | --- | --- |
| Pre-PCI model (model #1) | AKI | 9 | 0.8197 (0.8050 - 0.8344) | 0.7745 (0.7472 - 0.8019) |
|  | Bleeding | 11 | 0.8518 (0.8372 - 0.8645) | 0.8508 (0.8287 - 0.8728) |
|  | Stroke | 65 | 0.7266 (0.6574 - 0.7957) | 0.7174 (0.6163 - 0.8186) |
|  | Death | 8 | 0.9196 (0.9004 - 0.9386) | 0.9116 (0.8737 - 0.9494) |
|  | Any AO | 9 | 0.8266 (0.8154 - 0.8379) | 0.7981 (0.7782 - 0.8180) |
| PCI model (model #2) | AKI | 9 | 0.6579 (0.6390 - 0.6768) | 0.6253 (0.5923 - 0.6583) |
|  | Bleeding | 18 | 0.7745 (0.7563 - 0.7926) | 0.7349 (0.7031 - 0.7607) |
|  | Stroke | 49 | 0.5690 (0.4885 - 0.6495) | 0.4711 (0.3521 - 0.5900) |
|  | Death | 13 | 0.8429 (0.8149 - 0.8710) | 0.8126 (0.7618 - 0.8633) |
|  | Any AO | 14 | 0.7107 (0.6967 - 0.7248) | 0.6782 (0.6535 - 0.7026) |
| Final model | AKI | - | 0.8175 (0.8023 - 0.8326) | 0.7793 (0.7521 - 0.8065) |
|  | Bleeding | - | 0.8763 (0.8629 - 0.8898) | 0.8590 (0.8363 - 0.8818) |
|  | Stroke | - | 0.7293 (0.6594 - 0.7993) | 0.6990 (0.5882 - 0.8098) |
|  | Death | - | 0.9233 (0.9035 - 0.9431) | 0.9035 (0.8684 - 0.9386) |
|  | Any AO | - | 0.8372 (0.8261 - 0.8482) | 0.8057 (0.7862 - 0.8252) |

### Prediction of outcomes using pre-PCI variables

As shown in Table 2, Model 1 (pre-PCI probability of outcome) predicted AKI with an accuracy of 82% and 77% accuracy in the training and test sets, respectively. Similarly, high predictive accuracy was observed for bleeding (85% and 85%, respectively), death (92% and 91%, respectively) and any adverse outcome (83% and 80%, respectively). However, Model 1 had a suboptimal performance to predict stroke (73% and 72%, respectively). Concomitantly, the learning was quick for AKI, bleeding, death and any adverse outcome (optimum number of training epochs needed were only 8–9) as compared to that for stroke (65 epochs). The full model optimization and the mean squared error of prediction for all outcomes is shown in Supplementary Figure 1.

### Prediction of outcomes using PCI variables

The predictive performance of Model 2 (PCI-related probability of outcome) was, in general, less impressive than that of Model 1. This model provided optimum performance with one hidden layer containing 13 neurons. As shown in Table 2, the best predictions from Model 2 were for death (84% in training set, 81% in test set), bleeding (77% and 73%, respectively), any adverse outcome (71% and 68%, respectively) and AKI (66% and 63%, respectively). Learning took between 9 and 18 epochs for these outcomes. Consistently, the prediction of stroke was both poorer (57% and 47%, respectively) and slower (49 epochs).

### Prediction of outcomes using pre-PCI and PCI variables

The accuracy of the final model that combined predictions from both Model 1 and Model 2 is described in Table 2 and depicted in Figure 2 (in the test set only). In general, the predictive performance of the final model was marginally superior to that of Model 1 for all outcomes. In the training and test sets the predictive performance of the final model was best for death (92% and 90%, respectively), bleeding (88% and 86%, respectively), any adverse outcome (84% and 81%, respectively) and AKI (82% and 78%, respectively). Stroke could be predicted with accuracy of 73% and 70% in these datasets, respectively. Considering that the final predictive accuracy was primarily obtained from the pre-PCI probability (Model 1), we examined if specific subsets of patients with high or low likelihood of the event can be identified based on the probability estimates from Models 1 and 2. For this, we generated deciles of the predictions from Model 1 and 2, separately and estimated the likelihood ratio of the outcome for combinations of these deciles. The results are shown in Supplementary Figure 2. We found that higher deciles of the pre-PCI probability were associated with very high LRs for all outcomes. The association of the PCI-related probability (Model 2) deciles was, however, not linear for all outcomes. For instance, the LR for AKI was high (11.63) for patients in the highest decile of pre-PCI probability but in the lowest decile for PCI-related probability. Likewise, the LR for any adverse outcome was high (11.68) for patients in the 9th decile for pre-PCI probability but the lowest decile for PCI-related probability. Also, the LR for death was highest (62.1) for patients in the highest decile for pre-PCI probability but 6^th^ decile for PCI-related probability. These results indicated that the likelihood of study outcomes was primarily predicted by the pre-PCI probability and had complex, non-linear associations with the PCI-related probability of the event.

**Figure 2.**
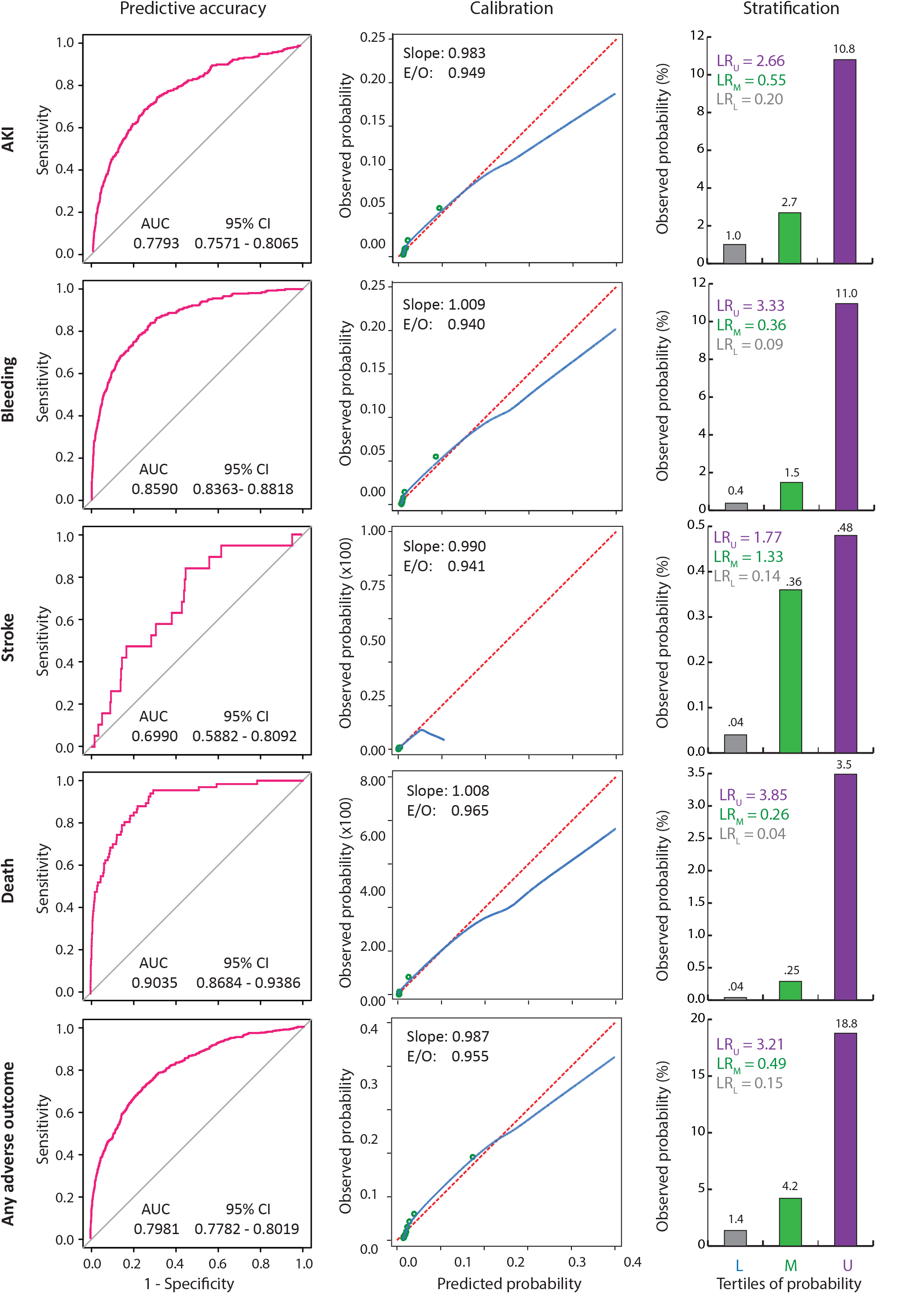
Predictive performance of the MLP-based classifier in the test set (n = 7,001). This trellis plot shows the accuracy (ROC curves), calibration (calibration plots) and stratification (bar graphs) for each outcome indicated on the left. AUC, area under the ROC curve; CI, confidence interval; E/O, expected over observed ratio; LR, level-specific likelihood ratio (subscripts indicate the tertiles); L, M, U, lower, middle and upper tertile, respectively.

### Predictive importance of input variables

In Supplementary Table 2, we list the top 20 important variables from Model 1 and top 10 important variables from Model 2 for each outcome. In general, the importance metrics for the variables included in Model 2 were lower than for those in Model 1 corroborating the observations that pre-PCI probability was a stronger predictor (than PCI-related probability) of the study outcomes. Of note, even though same set of inputs were provided to the MLP for each outcome, the set of important predictors varied by outcome. For AKI, not being on dialysis, having CKD, undergoing emergent PCI as an inpatient and the pre-PCI troponin T were the top five pre-PCI predictors. For bleeding, the top five pre-PCI predictors were history of other major surgery, anemia, cardiogenic shock and emergent or salvage PCI. For death, the top five pre-PCI predictors were pre-PCI cardiogenic shock (within 24 hours or 2 weeks), cardiac arrest, heart failure within two weeks and stenosis in the circumflex artery. For stroke, the importance metrics were not impressive but the top five pre-PCI variables included history of heart failure, stenosed ramus or proximal LAD in the absence of dyslipidemia and chronic lung disease. For any adverse outcome, the top five pre-PCI variables were a conglomerate of the above-mentioned variables for each outcome and included cardiogenic shock, history of major surgery, anemia, currently not on dialysis and an emergent PCI. Interestingly, across the outcomes, the important PCI-related variables primarily included insertion of intra-aortic balloon pump (IABP) or other mechanical ventricular support (MVS) before, during or after the PCI. Notably, nonuse of bleeding avoidance strategies (trans radial access, bivalirudin use and vascular closure device) was an important predictor of bleeding.

### Validation of the MLP classifier in the test set

The ROC curves for prediction of each outcome in the independent test set are shown in Figure 2 and the AUC for these curves is shown separately for each model in Table 2 (column titled “Accuracy in Test Set”). As mentioned above and shown in these ROC curves, the predictive performance in the test set was excellent for all outcomes except for stroke. The learning difficulty posed by the outcome of stroke was also exemplified further when we examined the calibration of the predicted probabilities from the final model (calibration plots in Figure 2). While all other outcomes had calibration slopes close to unity, the outcome of stroke had a low value for slope and a low range of predicted probabilities as indicated by a truncated calibration plot. The calibration slopes for death and bleeding were closest to unity, followed by any adverse outcome and AKI.

The predictive performance of the final model in the test set was typified by the appropriate risk stratification across tertiles of predicted probabilities for each study outcome. For example, the observed event rate of AKI in the low-, middle- and upper tertile of the predicted probability was 1%, 2.7% and 10.8%, respectively. Similar stratification was observed for the outcomes of bleeding (0.4%, 1.5% and 11%, respectively), death (0%, 0.3% and 3.5%, respectively), any adverse outcome (1.4%, 4.2% and 18.9%, respectively) and stroke (0.04%, 0.4% and 0.5%). Together, these results indicated, excellent accuracy, calibration and stratification provided by the final model for all outcomes other than stroke.

### Improvement in PCI risk-stratification by MLP model

Currently, the NCDR CathPCI models based on regression methods are used to encourage informed PCI decisions by providing pre-PCI estimates of predicted probability for death (19), bleeding (12) and AKI (20). We therefore compared the predicted probabilities obtained from the MLP models with these existing models to investigate whether the proposed method improves the predictive performance. The results of these analyses are shown in Table 3. We observed that for all three outcomes, the MLP based models provided significantly improved predictive accuracy (1.6%, 5.3% and 5.6% for death, bleeding and AKI, respectively), discrimination (6.0%, 7.2% and 4.9%, respectively) and reclassification (continuous NRI 0.61, 0.18, 0.07, respectively). Together, these results demonstrate the improved predictive performance using the perceptron-based models over the existing methods.

**Table 3.** Improved risk-stratification using the MLP-based classification

| Characteristic | Parameter | AKI | Bleeding | Death |
| --- | --- | --- | --- | --- |
| Accuracy | $\Delta\text{AUC}$, p | 0.0564, $2.2 \times 10^{-26}$ | 0.0526, $4.4 \times 10^{-23}$ | 0.0161, 0.0489 |
| Discrimination | IDI, p | 0.0486, $4.0 \times 10^{-31}$ | 0.0717, $9.2 \times 10^{-34}$ | 0.0602, $4.7 \times 10^{-7}$ |
| Classification | NRI, p | 0.0729, 0.0110 | 0.1765, $3.3 \times 10^{-8}$ | 0.6078, $1.4 \times 10^{-25}$ |

## DISCUSSION

We have developed a system of models using the ANN framework and MLP structure to simultaneously predict three PCI-related adverse outcomes – bleeding, AKI and death. A comparison of our predictive models with the current standard-of-care predicting models showed a significant improvement in accuracy, discrimination and reclassification. Recently, Huang et al (21) published an elegant model to predict AKI after PCI using the large NCDR CathPCI dataset emanating from 1,694 US hospitals. They employed a generalized additive model to account for the potential nonlinear relationships. Depending on the definition of AKI (degree of rise in serum creatinine post-PCI) their model accuracy ranged from 77.7% to 84.9% with the more severe forms of AKI (serum creatinine absolute rise > 1.0 mg/dl over baseline) predicted more accurately. Our definition of AKI corresponded to a mild increase in serum creatinine (absolute rise > 0.3 mg/dl). The predictive accuracy of our model for AKI (77.9%) was therefore comparable with that of the model developed by Huang et al.(21) In a similar fashion, risk models for predicting bleeding after PCI have traditionally reported predictive accuracy in the range of 65% to 79%.(22–24) A meta-analysis of 6 published studies that included separate validation datasets found that the aggregate predictive accuracy of risk models for PCI-related bleeding was 68% (95% CI 65% – 72%).(25) Thus, the current models for prediction of bleeding demonstrate only moderate level of accuracy. In contrast, our ANN-based model predicted bleeding with 86.5% accuracy – a substantial improvement over current models. The best predictive performance of our model, however, was for the outcome of all-cause mortality during hospitalization (predictive accuracy 90.3%) and was comparable with that of the currently used models (3) for mortality prediction. Together, our system of ANN models offered an improved and accurate prediction of three, PCI-related outcomes during index hospitalization – AKI, bleeding and death.

We and others have demonstrated the importance of contrast use limitation as a predictor of post-PCI AKI. Out second stage model for AKI did not include contrast use in the top 10 contributors to AKI prediction, neither did the S2 model greatly increase the performance of the stage 1 model (data shown in Table 2 and Supplementary Table 2). This result implies that in the dataset studied, the likelihood of contrast induced AKI may have been limited. In general, for all outcomes the stage 2 models in our dataset made only a small contribution to the final predictive accuracy. Training the models on larger datasets with richer procedural characteristics can therefore be expected to further augment the predictive performance of all the models.

Our model did not provide a satisfactory prediction of stroke after PCI. In this context, it is noteworthy that while efforts are underway to understand the determinants of stroke after PCI, currently there are no risk prediction models for this outcome. Hence comparison of the predictive performance of our model for stroke with other models is not possible. It is understood that trans radial intervention(26), receipt of ventilation, circulatory support, thrombolysis and acute coronary syndrome are predictors of ischemic or hemorrhagic stroke after PCI.(27) The list of important variables in our model (Supplementary Table 2) only partially concurred with this list. The inability of our model to predict stroke after PCI can be explained in part on the basis of following three reasons. First, the outcome of stroke in our study represented a composite outcome that included TIA, ischemic stroke and hemorrhagic stroke. As shown by Myint et al,(27) determinant of differ types of stroke after PCI may be different. Second, the prevalence of stroke in our dataset was low (0.27%) representing a scenario of potential imbalanced machine learning. A possible lack of adequate number of training sample in the stroke class could have limited the predictive ability of our model. Third, stroke maybe an inherently harder outcome to predict as compared to AKI, bleeding and death. Larger datasets with adequate number of training examples are needed before our approach can be used for prediction of stroke after PCI.

Our study used observational data and thus all the limitations implicit in such a dataset can be expected to be operational in our study as well. In addition, following limitations need to be considered. First, while all the models are temporally valid, the models can be made further clinically applicable by predicting time to events. Second, our model predicted the “any adverse outcome” as a composite outcome measure with good accuracy but a more specific model can be envisioned that can exploit larger datasets to predict specific combinations of PCI-related outcomes. Third, in line with the model by Huang et al,(21) it could be clinically more useful to test the model performance for varying levels of absolute rise in serum creatinine. However, in our dataset the prevalence of high absolute serum creatinine rise (>1mg/dl) was below 1% and thus difficult to predict. Larger datasets with adequate representation of AKI severity classes can be used in future studies to further refine the AKI component of our system of models.

In conclusion, we have developed a system of ANN-based predictive models that accurately predict three PCI-related outcomes – AKI, bleeding and death. In the era of personalized medicine, expensive interventions and bundled payments, this work has important implications. The direct impact of these improved ANN-based prediction of PCI-related outcomes needs to be investigated and determined in future studies.

## Data Availability

The code is available with the authors and can be shared upon reasonable request. The data is the property of the Barnes Jewish Hospital and cannot be shared.

